# Electrolyte Imbalance in Asphyxiated Term Neonates: Incidence, Predictors, and Outcomes from a Prospective Cohort study in Northern Uganda

**DOI:** 10.1101/2025.10.29.25339105

**Authors:** Bahari Yusuf, Tom Ediamu, Simon Odoch, Zeinab issa Ali, Yasin Ahmed H. Abshir, Abdulahi Abdirizak Farah, Abishir Mohamud Hersi, Ahmed Owjama, Hamdi M. Yusuf, Zakaria Abdi Said, Theoneste Hakizimana, Walyedin Elfakey, Grace Ndeezi

**Author notes:** Corresponding Author **Bahari Yusuf,** Email: address.

## Abstract

**Background:** Birth asphyxia is a major cause of neonatal morbidity and mortality, particularly in low-resource settings. Hypoxia and metabolic derangements that occur during asphyxia predispose neonates to electrolyte abnormalities, which may worsen the clinical course and contribute to poor outcomes. Early recognition and management of such imbalances can improve survival and prevent long-term neurological damage. This study aimed to determine the incidence, Predictors, and early outcomes of electrolyte imbalance among term neonates admitted with birth asphyxia in Lira Regional Referral Hospital, Uganda.

**Methods:** A hospital-based prospective cohort study was conducted among term neonates admitted with birth asphyxia, defined as a 5-minute Apgar score <7. Serum sodium, potassium, and calcium levels were measured at admission, and repeated on days 3, 7, and 14 for those still admitted. Clinical information including maternal and perinatal characteristics was recorded. Modified Poisson regression using SPSS was performed to identify independent predictors of electrolyte imbalance. Early outcomes, including mortality and length of hospital stay, were documented within the first 14 days.

**Results:** A total of 152 neonates were enrolled; 52.6% were male. During follow-up, 42 (29.0%) developed hyponatremia, 29 (19.5%) hyperkalemia, and 31 (21.2%) hypocalcemia. Independent predictors of hyponatremia included low Apgar score (0–3), severe hypoxic ischemic encephalopathy (HIE), prolonged intravenous fluid administration (>48 hours), resuscitation at birth, and dehydration. Convulsions, prolonged intravenous fluids, and resuscitation were significantly associated with hypocalcemia, while hyperkalemia was linked to low birth weight and prolonged intravenous fluids (p<0.05 for all). Mortality was highest among neonates with hyperkalemia, whereas hypocalcemia was significantly associated with prolonged hospital stay (p<0.05).

**Conclusion:** Electrolyte imbalances are common among term neonates with birth asphyxia, with hyponatremia, hypocalcemia, and hyperkalemia occurring in nearly one-fifth to one-third of cases. Routine electrolyte monitoring is essential in the management of asphyxiated neonates. Protocols for intravenous fluid use and timely correction of electrolyte derangements should be prioritized to reduce mortality and improve outcomes.

## Background

Birth asphyxia is one of the most serious neonatal emergencies, contributing substantially to global morbidity and mortality in the early neonatal period. It is defined by the World Health Organization as the inability to initiate and sustain breathing at birth, and is clinically identified using a 5-minute Apgar score of less than seven^1^. Globally, birth asphyxia affects between 2–9 neonates per 1,000 live births, with an estimated 4–9 million cases reported annually. It is responsible for approximately 900,000 neonatal deaths worldwide each year and accounts for nearly one quarter of neonatal mortality^2, 3^. The burden is disproportionately higher in low- and middle-income countries (LMICs), particularly in Sub-Saharan Africa and South Asia, where more than 98% of neonatal deaths occur^4^. In Uganda, birth asphyxia remains the leading cause of neonatal mortality, contributing to nearly 59% of neonatal deaths^3^.

The pathophysiological consequences of birth asphyxia are profound and multifactorial. Hypoxia and ischemia impair oxidative metabolism, leading to accumulation of lactic acid, energy failure, and dysfunction of cellular ion pumps. This cascade contributes to hypoxic– ischemic encephalopathy (HIE), renal impairment, myocardial dysfunction, and metabolic derangements^16, 18^. A critical yet often under-recognized complication in this context is electrolyte imbalance, particularly involving sodium, potassium, and calcium. These electrolytes play essential roles in neuronal excitability, cardiac function, neuromuscular stability, and fluid homeostasis. Even subtle derangements can precipitate seizures, arrhythmias, poor perfusion, or prolonged recovery in vulnerable neonates^5, 8, 9^.

Hyponatremia in asphyxiated neonates is commonly attributed to inappropriate secretion of antidiuretic hormone and impaired renal tubular reabsorption of sodium. It worsens cerebral edema, increases the risk of convulsions, and is associated with higher stages of HIE^7, 11^. Hyperkalemia arises from a shift of potassium from the intracellular to extracellular space due to cell membrane dysfunction, compounded by impaired renal excretion secondary to acute kidney injury. This imbalance significantly increases the risk of fatal cardiac arrhythmias^9, 15^. Hypocalcemia is believed to result from suppression of parathyroid hormone secretion and disrupted placental calcium transfer, and is strongly linked to hypotonia, cardiac dysfunction, and seizures^5, 8^. Together, these derangements amplify the vulnerability of asphyxiated neonates, compounding the effects of hypoxia and ischemia.

Evidence from South Asia and the Middle East highlights the high prevalence of these electrolyte imbalances among neonates with asphyxia. Studies in India, Bangladesh, Iraq, and Nepal have consistently reported that hyponatremia, hypocalcemia, and hyperkalemia are more common in neonates with severe HIE compared to those with mild or moderate disease^11–14^. For instance, one study in Nepal reported hyponatremia in 81.8%, hyperkalemia in 90.9%, and hypocalcemia in 45.4% of neonates with severe asphyxia^11^. Similar findings from Bangladesh and India demonstrate that electrolyte abnormalities are not only frequent but also strongly associated with increased mortality and prolonged hospitalization^12–15^. Despite this evidence, data from Sub-Saharan Africa remain sparse, and no study from Uganda had comprehensively examined the incidence, Predictors, and outcomes of these derangements among asphyxiated neonates^6, 17^.

In Uganda, routine neonatal care protocols for birth asphyxia largely focus on stabilization, resuscitation, and supportive management. Serum electrolyte monitoring is not consistently integrated into the management of neonates admitted with birth asphyxia, largely due to resource constraints^3^. Observations at Lira Regional Referral Hospital revealed that neonates admitted with low Apgar scores often presented with refractory seizures, severe shock, or prolonged hospital stays, yet electrolyte disturbances were not routinely investigated or corrected. This gap in clinical practice highlights the urgent need for local data to inform guidelines, optimize care, and improve neonatal survival^17^.

This study was therefore conducted to determine the incidence, Predictors, and early outcomes of electrolyte imbalance among term neonates admitted with birth asphyxia at Lira Regional Referral Hospital in Northern Uganda. The findings are expected to provide vital evidence to guide routine monitoring, inform neonatal management protocols, and contribute to strategies aimed at reducing neonatal morbidity and mortality in resource-limited settings.

## Methods

### Study design and setting

This was a hospital-based prospective cohort study carried out in the Neonatal Intensive Care Unit (NICU) of Lira Regional Referral Hospital (LRRH) in Northern Uganda. LRRH is a major public referral facility serving districts in the Lango sub-region, including Lira, Amolatar, Apac, Dokolo, Kole, and Oyam. It is located approximately 339 kilometers north of Kampala, Uganda’s capital. The NICU at LRRH receives a large number of neonates admitted with low Apgar scores and birth asphyxia every month, making it an appropriate setting for the present study. Data collection was conducted over a period of three months(March-july 2025) following ethical approval.

### Study population

The study population consisted of term neonates admitted to the NICU of LRRH with a clinical diagnosis of birth asphyxia. Birth asphyxia was defined as a 5-minute Apgar score of less than seven. Only neonates who were born at a gestational age of 37 weeks or more and who met the diagnostic criteria for asphyxia were considered eligible for participation.

### Eligibility criteria

Neonates were eligible if they were born at term, admitted to the NICU with a diagnosis of birth asphyxia, and if their parents or guardians provided written informed consent. Neonates with major congenital malformations or those who were born preterm were excluded from the study.

### Sample size determination

For objective one, the sample size was estimated using Daniel’s formula *n = z^2^pq/d^2^*. Using findings from Bangladesh where 10% of neonates with asphyxia had hyperkalemia^19^, with *p = 0.10*, *d = 0.05*, and *z = 1.96* at 95% confidence, the calculated sample size was 138.

For objective two, the Epi-Info online calculator (Fleiss method) was applied^20^ using results from India, where electrolyte abnormalities increased with HIE stage (46.6% in stage I and 71.4% in stage III)^21^. At 80% power and 95% confidence, the required sample was 122.

For objective three, the Epi-Info calculator (Fleiss method) was used^20^ with data from Bangladesh showing 60% mortality among neonates with hypokalemia versus 22.6% with normal potassium^15^. This gave a sample size of 64, which rose to 71 after adding 10% for loss to follow-up. Taking the largest estimate (138) and adding 10% for attrition, the final sample size was 152 neonates. Consecutive enrolment was employed until the required sample was attained.

### Study procedure

Upon admission to the NICU, neonates were assessed for eligibility. After obtaining informed consent from the parents or guardians, baseline demographic information and perinatal history were collected using a structured questionnaire. Clinical evaluation was then performed, which included assessment of vital signs, staging of hypoxic ischemic encephalopathy (HIE) using Thomson score, and documentation of complications such as seizures, respiratory distress, shock, or dehydration. Anthropometric measurements including birth weight and length were taken at the time of enrollment.

Blood samples were obtained for serum electrolyte estimation at admission. Neonates with normal electrolytes at baseline were followed up during hospitalization, and repeat measurements were performed on day 3, day 7, and day 14 for those who remained admitted. Clinical progress was monitored daily, focusing on convulsions, urine output, dehydration status, and intravenous fluid administration. Outcomes were recorded within the first 14 days of admission, including survival, death, and length of hospital stay. A prolonged stay was defined as more than 10 days, corresponding to the 75th percentile of the data.

### Laboratory procedure

Approximately 2 to 3 mL of venous blood was collected under aseptic precautions into plain vacutainer tubes. Samples were allowed to clot and centrifuged to separate serum. Sodium and potassium levels were measured using the ion-selective electrode method, while total calcium levels were measured using the Arsenazo III colorimetric method. Serum albumin was also assayed, and corrected calcium was calculated accordingly to account for hypoalbuminemia. The analysis was carried out in the LRRH central laboratory, which routinely undertakes internal quality control procedures. Normal ranges were defined as sodium 135–145 mEq/L, potassium 3.5–5.5 mEq/L, and calcium 7.6–11.3 mg/dL. Any value outside these ranges was classified as an electrolyte imbalance. To ensure accuracy, equipment was calibrated daily, and 10% of randomly selected samples were re-tested to confirm consistency of results.

### Data management and analysis

Data were initially entered into Microsoft Excel, cleaned, and then exported to SPSS version 25 for statistical analysis. Descriptive statistics were computed to summarize baseline characteristics and the incidence of electrolyte imbalances. Bivariate analysis was conducted to assess associations between potential Predictors and electrolyte disturbances. Variables with a p-value less than 0.2 were included in multivariate analysis using modified Poisson regression with robust error variance to identify independent predictors. The strength of associations was reported using adjusted risk ratios with 95% confidence intervals, and statistical significance was set at a p-value less than 0.05.

### Ethical considerations

Ethical approval for the study was obtained from the Kampala International University Research and Ethics Committee (KIU-REC). Administrative clearance was granted by the management of LRRH. Informed consent was obtained from all parents or guardians of eligible neonates prior to enrollment. Confidentiality was maintained by de-identifying participant information, and access to data was restricted to the research team. All neonates identified with electrolyte imbalances received appropriate management in accordance with hospital protocols.

## Results

### Baseline characteristics

A total of 152 neonates with birth asphyxia were enrolled. Slightly over half were male (80/152, 52.6%), and 67.8% were admitted <6 hours after birth (103/152). Most had an Apgar score 4–6 (138/152, 90.8%), with 9.2% scoring 0–3. Low birth weight (<2,500 g) occurred in 9.2% (14/152), while 90.8% weighed 2,500–4,000 g. Resuscitation was required in 91.4% (139/152); recorded resuscitation duration was 1–5 min in 23.7%, 6–10 min in 21.7%, and >10 min in 46.1%. By HIE severity, mild 61.2% (93/152), moderate 21.7% (33/152), and severe 17.1% (26/152). Respiratory distress was present in 57.2% (87/152), dehydration in 12.5% (19/152), neonatal sepsis in 25.7% (39/152), and convulsions in 46.1% (70/152). Anuria was uncommon (no urine passed in 3.9%, 6/152), and shock was documented in 3.9% (6/152). For IV fluids, the recorded duration was ≤48 hours in 42.1% and >48 hours in 57.9%. At admission, electrolytes were mostly normal: sodium normal 95.4% (145/152), potassium normal 98.0% (149/152), and calcium normal 96.1% (146/152). The baseline clinical and demographic characteristics of the study participants are summarized in **Table 1**.

**Table 1:**
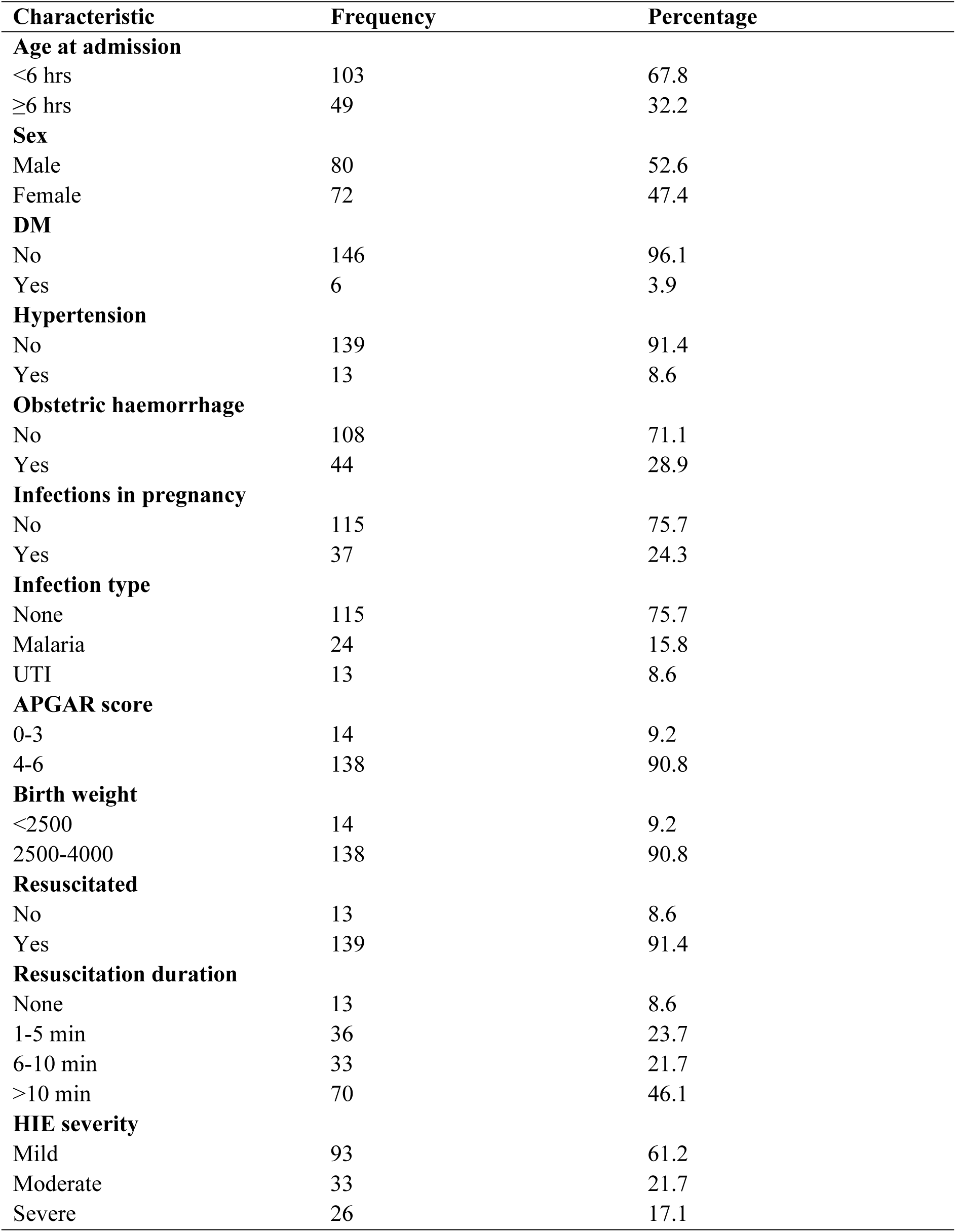
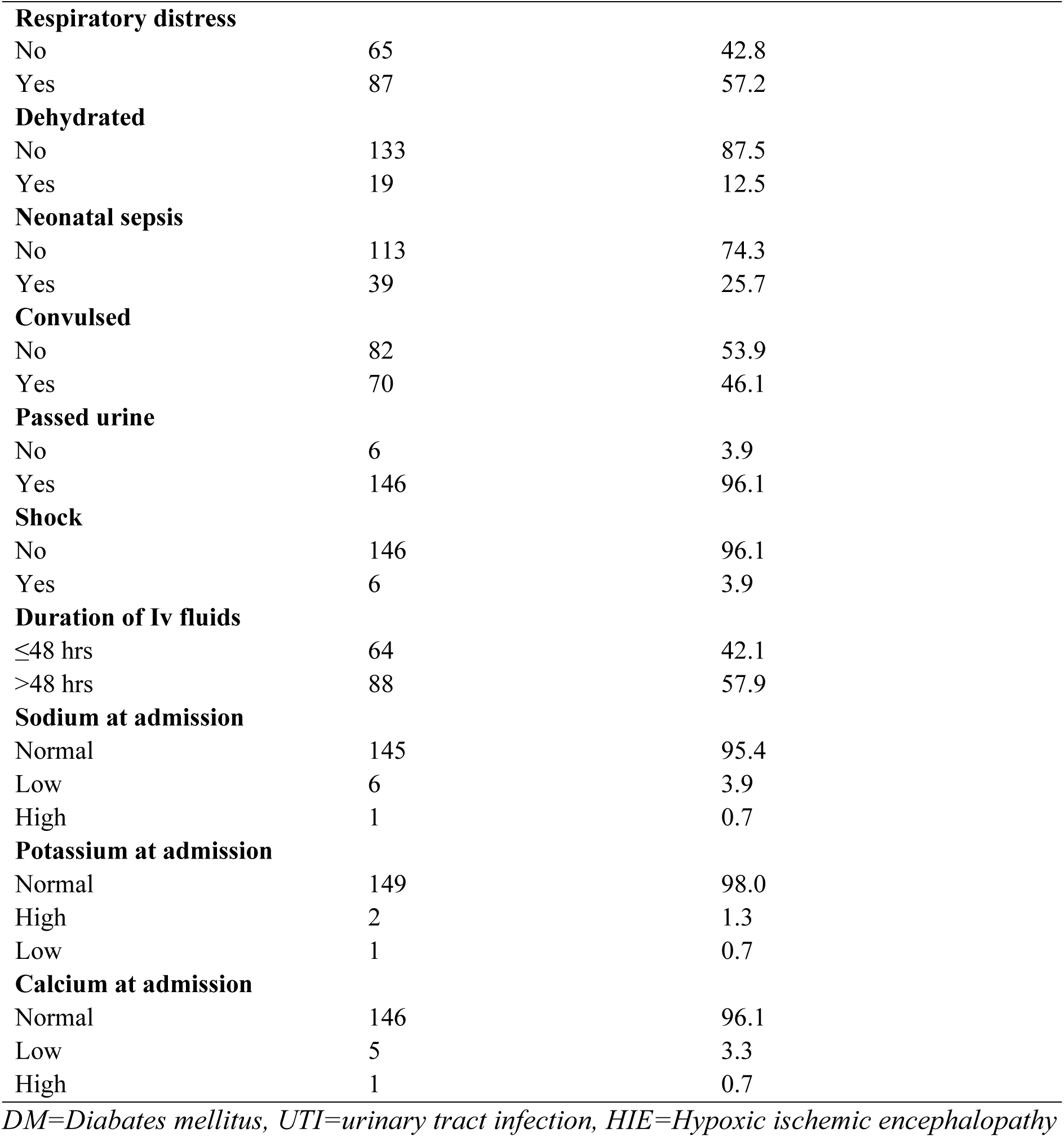
Baseline characteristics of study participants.

| Characteristic | Frequency | Percentage |
| --- | --- | --- |
| **Age at admission** |  |  |
| <6 hrs | 103 | 67.8 |
| ≥6 hrs | 49 | 32.2 |
| **Sex** |  |  |
| Male | 80 | 52.6 |
| Female | 72 | 47.4 |
| **DM** |  |  |
| No | 146 | 96.1 |
| Yes | 6 | 3.9 |
| **Hypertension** |  |  |
| No | 139 | 91.4 |
| Yes | 13 | 8.6 |
| **Obstetric haemorrhage** |  |  |
| No | 108 | 71.1 |
| Yes | 44 | 28.9 |
| **Infections in pregnancy** |  |  |
| No | 115 | 75.7 |
| Yes | 37 | 24.3 |
| **Infection type** |  |  |
| None | 115 | 75.7 |
| Malaria | 24 | 15.8 |
| UTI | 13 | 8.6 |
| **APGAR score** |  |  |
| 0-3 | 14 | 9.2 |
| 4-6 | 138 | 90.8 |
| **Birth weight** |  |  |
| <2500 | 14 | 9.2 |
| 2500-4000 | 138 | 90.8 |
| **Resuscitated** |  |  |
| No | 13 | 8.6 |
| Yes | 139 | 91.4 |
| **Resuscitation duration** |  |  |
| None | 13 | 8.6 |
| 1-5 min | 36 | 23.7 |
| 6-10 min | 33 | 21.7 |
| >10 min | 70 | 46.1 |
| **HIE severity** |  |  |
| Mild | 93 | 61.2 |
| Moderate | 33 | 21.7 |
| Severe | 26 | 17.1 |
| **Respiratory distress** |  |  |
| No | 65 | 42.8 |
| Yes | 87 | 57.2 |
| **Dehydrated** |  |  |
| No | 133 | 87.5 |
| Yes | 19 | 12.5 |
| **Neonatal sepsis** |  |  |
| No | 113 | 74.3 |
| Yes | 39 | 25.7 |
| **Convulsed** |  |  |
| No | 82 | 53.9 |
| Yes | 70 | 46.1 |
| **Passed urine** |  |  |
| No | 6 | 3.9 |
| Yes | 146 | 96.1 |
| **Shock** |  |  |
| No | 146 | 96.1 |
| Yes | 6 | 3.9 |
| **Duration of Iv fluids** |  |  |
| ≤48 hrs | 64 | 42.1 |
| >48 hrs | 88 | 57.9 |
| **Sodium at admission** |  |  |
| Normal | 145 | 95.4 |
| Low | 6 | 3.9 |
| High | 1 | 0.7 |
| **Potassium at admission** |  |  |
| Normal | 149 | 98.0 |
| High | 2 | 1.3 |
| Low | 1 | 0.7 |
| **Calcium at admission** |  |  |
| Normal | 146 | 96.1 |
| Low | 5 | 3.3 |
| High | 1 | 0.7 |

*DM=Diabetes mellitus, UTI=urinary tract infection, HIE=Hypoxic ischemic encephalopathy*

### Incidence of electrolyte imbalance

At baseline, the majority of neonates had normal electrolyte levels and were followed during hospitalization. For sodium, 145 neonates had normal values at admission, of whom 42 (29.0%; 95% CI 22.1–36.6) developed hyponatremia during follow-up, with most cases detected on day 3. For potassium, 149 neonates had normal levels initially, and 29 (19.5%; 95% CI 13.4–26.2) developed hyperkalemia, again predominantly identified on day 3. For calcium, 146 neonates were normal at baseline, and 31 (21.2%; 95% CI 14.4–28.8) developed hypocalcemia, the majority also first noted on day 3. Overall, these findings demonstrate that electrolyte disturbances are common among asphyxiated neonates, often emerging within the first few days of life, underscoring the need for routine monitoring during this critical period. The incidence of electrolyte imbalance among term neonates with birth asphyxia is presented in **Table 2** below.

**Table 2:**
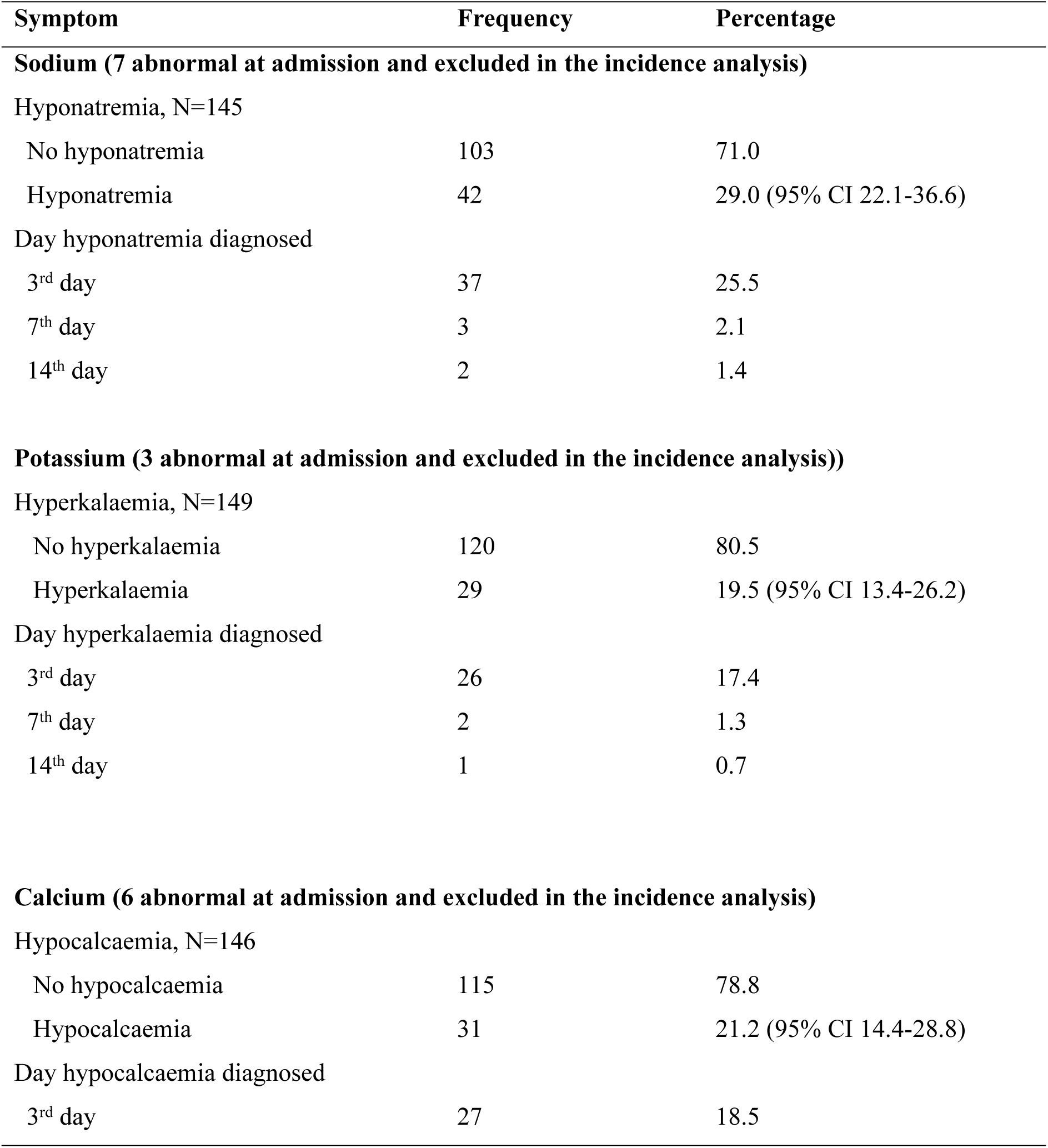
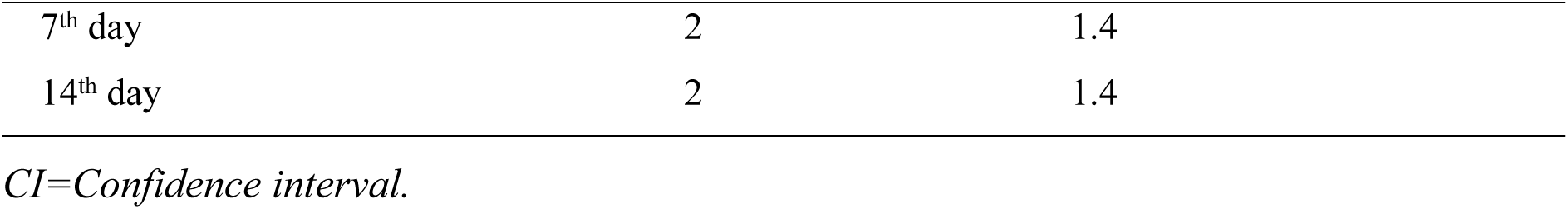
Incidence of electrolyte imbalance among term neonates admitted with birth.

### Predictors for Electrolyte Imbalance

In the multivariable analysis, several independent predictors of hyponatremia were identified. Neonates with a very low Apgar score (0–3) at five minutes were 54.9% more likely to develop hyponatremia compared to those with higher scores. Resuscitation at birth increased the risk by 22.7%, while severe hypoxic ischemic encephalopathy (HIE) raised the risk by 32.1%. Dehydration was a particularly strong predictor, increasing the likelihood of hyponatremia by 62.7%. Prolonged intravenous fluid administration beyond 48 hours was also significantly associated with hyponatremia, raising the risk by 28.5%.

For hyperkalemia, the independent predictors were low birth weight and prolonged intravenous fluid use. Neonates with a birth weight below 2,500 g had a 35.1% higher risk of developing hyperkalemia, while those who received intravenous fluids for more than 48 hours had a 25.0% increased risk. The majority of neonates who developed hyperkalemia (89.7%) had received intravenous fluids for over 48 hours.

Hypocalcemia was independently associated with resuscitation at birth, convulsions, and prolonged intravenous fluid therapy. Neonates who were resuscitated had a 19.6% higher risk, those with convulsions had a 16.9% increased risk, and those who received intravenous fluids beyond 48 hours had a 28.4% increased risk of developing hypocalcemia. A large proportion of affected neonates had undergone resuscitation (100%) and received prolonged intravenous fluids (77.4%). Independent predictors of hyponatremia, hyperkalemia, and hypocalcemia are summarized in **Tables 3, 4, and 5,** respectively.

**Table 3:** Multivariable analysis of Predictors of hyponatremia among term neonates admitted with birth asphyxia.

| Characteristic | Bivariable analysis |  |  | Multivariable analysis |  |  |
| --- | --- | --- | --- | --- | --- | --- |
|  | cRR | 95% CI | P value | aRR | 95% CI | P value |
| APGAR score |  |  |  |  |  |  |
| 0-3 | 1.874 | 1.539-2.282 | <0.001 | 1.549 | 1.201-1.999 | 0.001* |
| 4-6 | Ref |  |  |  |  |  |
| Resuscitated |  |  |  |  |  |  |
| No | Ref |  |  |  |  |  |
| Yes | 1.263 | 1.071-1.490 | 0.005 | 1.227 | 1.030-1.463 | 0.022* |
| HIE severity |  |  |  |  |  |  |
| Mild | Ref |  |  |  |  |  |
| Moderate | 1.140 | 0.955-1.361 | 0.146 | 1.049 | 0.895-1.229 | 0.552 |
| Severe | 1.771 | 1.469-2.135 | <0.001 | 1.321 | 1.078-1.620 | 0.007* |
| Dehydrated |  |  |  |  |  |  |
| No | Ref |  |  |  |  |  |
| Yes | 1.713 | 1.382-2.122 | <0.001 | 1.627 | 1.280-2.066 | <0.001 |
| Duration of IV fluids |  |  |  |  |  |  |
| ≤48 hrs | Ref |  |  |  |  |  |
| >48 hrs | 1.315 | 1.149-1.505 | <0.001 | 1.285 | 1.119-1.495 | 0.001* |

*cRR= Crude rate ratio, aRR= adjusted rate ratio, CI= Confidence interval.*

**Table 4:** Multivariable analysis of Predictors of hyperkalemia among term neonates admitted with birth asphyxia.

| Characteristic | Bivariable analysis |  |  | Multivariable analysis |  |  |
| --- | --- | --- | --- | --- | --- | --- |
|  | cRR | 95% CI | P value | aRR | 95% CI | P value |
| Obstetric haemorrhage |  |  |  |  |  |  |
| No | Ref |  |  |  |  |  |
| Yes | 1.202 | 1.030-1.404 | 0.020 | 1.140 | 0.994-1.309 | 0.062 |
| APGAR score |  |  |  |  |  |  |
| 0-3 | 1.295 | 0.991-1.691 | 0.058 | 1.297 | 0.962-1.749 | 0.087 |
| 4-6 | Ref |  |  |  |  |  |
| Birth weight |  |  |  |  |  |  |
| <2500 | 1.401 | 1.070-1.834 | 0.014 | 1.351 | 1.047-1.743 | 0.021* |
| 2500-4000 | Ref |  |  |  |  |  |
| Duration of IV fluids |  |  |  |  |  |  |
| ≤48 hrs | Ref |  |  |  |  |  |
| >48 hrs | 1.279 | 1.146-1.428 | <0.001 | 1.250 | 1.079-1.448 | 0.003* |

*cRR= Crude rate ratio, aRR= adjusted rate ratio, CI= Confidence interval.*

**Table 5:** Multivariable analysis of Predictors of hypocalcemia among term neonates admitted with birth asphyxia.

| Characteristic | Bivariable analysis |  |  | Multivariable analysis |  |  |
| --- | --- | --- | --- | --- | --- | --- |
|  | cRR | 95% CI | P value | aRR | 95% CI | P value |
| Resuscitated |  |  |  |  |  |  |
| No | Ref |  |  |  |  |  |
| Yes | 1.262 | 1.175-1.357 | <0.001 | 1.196 | 1.034-1.383 | 0.016* |
| Convulsed |  |  |  |  |  |  |
| No | Ref |  |  |  |  |  |
| Yes | 1.280 | 1.032-1.349 | 0.016 | 1.169 | 1.021-1.337 | 0.026* |
| Duration of Iv fluids |  |  |  |  |  |  |
| ≤48 hrs | Ref |  |  |  |  |  |
| >48 hrs | 1.195 | 1.055-1.353 | 0.005 | 1.284 | 1.106-1.490 | 0.006* |

*cRR= Crude rate ratio, aRR= adjusted rate ratio, CI= Confidence interval.*

### Early Outcomes of Electrolyte Imbalance

Overall mortality was 11.8% (18 of 152 neonates). Although mortality was higher across all electrolyte derangements, the difference was only statistically significant for hyperkalemia. Among the 29 neonates who developed hyperkalemia, 12 (41.4%) died within 14 days, compared with 4 of 120 (3.3%) neonates without hyperkalemia (p<0.001). Hyponatremia was not significantly associated with mortality: 6 of 42 neonates with hyponatremia (14.3%) died compared to 11 of 103 (10.7%) who remained normonatremic (p=0.574). Similarly, hypocalcemia showed no statistically significant association with mortality, as 6 of 31 affected neonates (19.4%) died compared with 12 of 115 (10.4%) without hypocalcemia (p=0.217). below is **Table 6** which summarizes mortality outcomes.

**Table 6:**
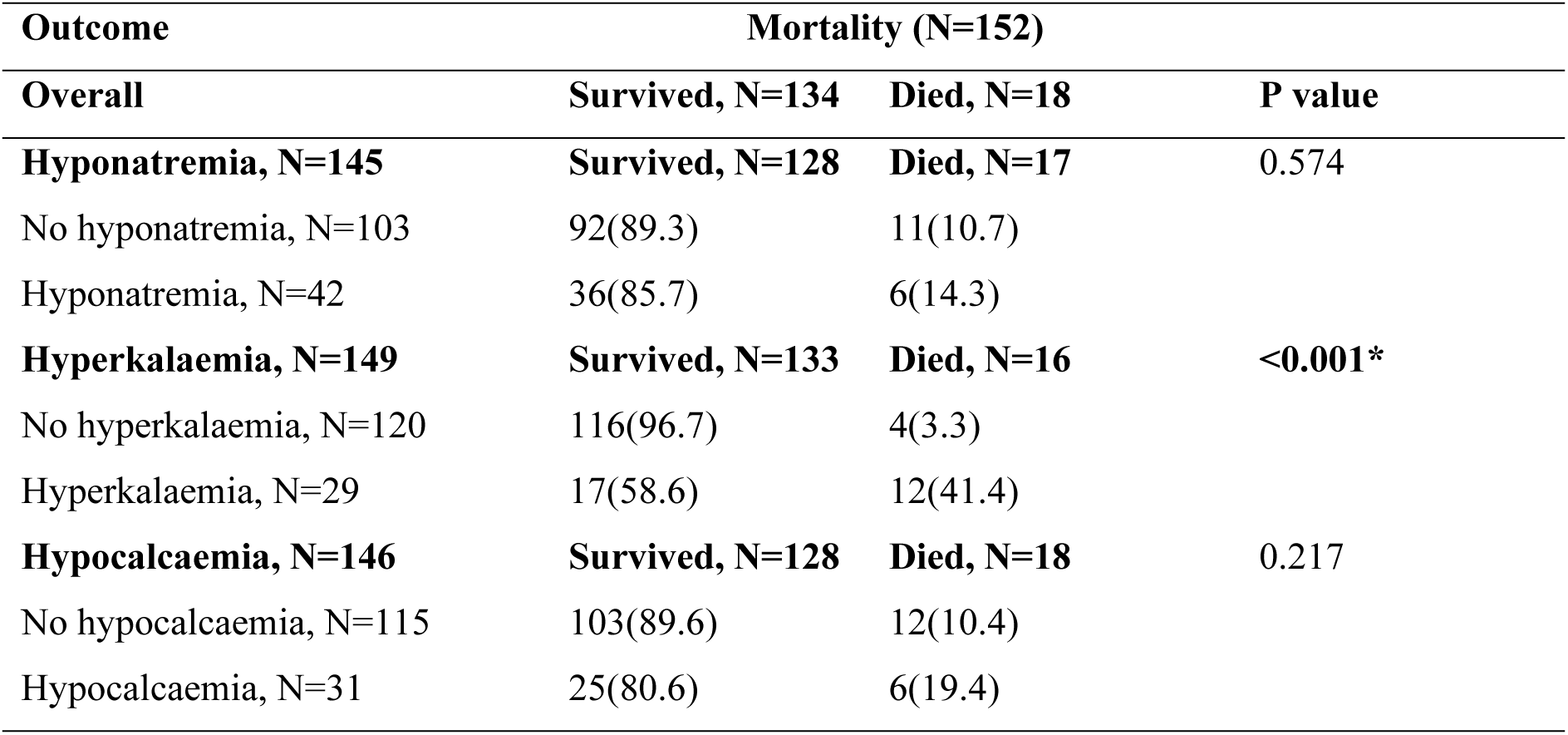
Mortality associated with electrolyte imbalance among term neonates admitted with birth asphyxia.

The proportion of neonates with prolonged hospital stay was higher among those with electrolyte disturbances, but the difference was statistically significant only for hypocalcemia. Of the 31 neonates who developed hypocalcemia, 15 (48.4%) had prolonged hospital stay beyond 10 days compared to 18 of 97 (18.6%) among those without hypocalcemia (p=0.002). Neither hyponatremia nor hyperkalemia showed a significant association with prolonged hospital stay. **Table 7** below presents the relationship between electrolyte imbalance and hospital stay duration.

**Table 7:**
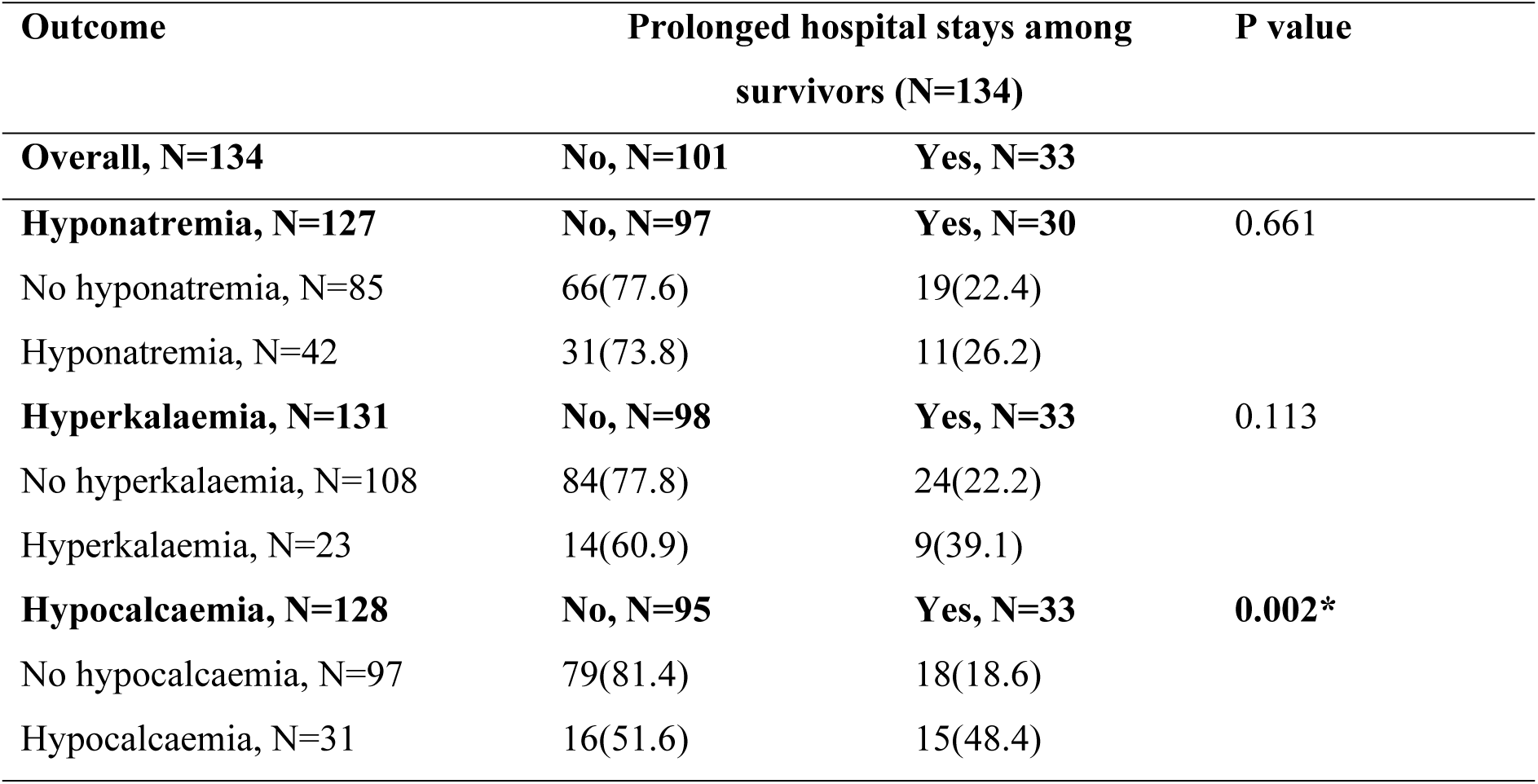
Hospital stay in relation to electrolyte imbalance among term neonates admitted with birth asphyxia.

## Discussion

This prospective study demonstrated a high incidence of electrolyte imbalance among term neonates admitted with birth asphyxia at Lira Regional Referral Hospital. Hyponatremia was observed in 29.0% of neonates, hyperkalemia in 19.5%, and hypocalcemia in 21.2%. Most of these disturbances emerged by day 3 of admission, underscoring the importance of early monitoring. The incidence observed in our study is consistent with reports from South Asia and the Middle East, where electrolyte abnormalities are common among asphyxiated neonates^11–15^. For example, studies in Nepal and Bangladesh reported hyponatremia ranging between 28–42% and hypocalcemia in 20–40% of cases, figures that are closely aligned with our findings^11, 12^. However, our incidence of hyperkalemia (19.5%) was lower than that reported in some Asian cohorts, where rates as high as 45–90% have been documented^11, 13, 15^. This discrepancy may reflect differences in disease severity, timing of blood sampling, and local NICU practices such as fluid administration.

In the analysis of Predictors, we found that hyponatremia was significantly associated with severe HIE, very low Apgar scores (0–3), dehydration, convulsions, resuscitation at birth, and prolonged intravenous fluid administration. These associations are biologically plausible, as hypoxia impairs renal tubular function and stimulates antidiuretic hormone secretion, leading to water retention and dilutional hyponatremia^7, 11^. Our findings are in agreement with studies from India and Pakistan, which demonstrated that hyponatremia correlated with the severity of HIE and was more common in neonates who required resuscitation^12, 13^. Similar associations have been observed in Iraq, where dehydration and seizures were strong predictors of sodium imbalance^13^.

Hyperkalemia in our cohort was independently associated with low birth weight and prolonged intravenous fluid therapy. These results are consistent with earlier work from Bangladesh and Nepal, where low birth weight neonates were particularly vulnerable to potassium disturbances, partly due to renal immaturity and increased cellular breakdown^11, 12, 15^. The link with prolonged intravenous fluids likely reflects impaired excretion and dilutional effects. Importantly, the majority of deaths in our study occurred among neonates with hyperkalemia, reinforcing the clinical significance of this finding. Other studies, including those from India and Bangladesh, have similarly reported that hyperkalemia is the most lethal electrolyte imbalance in birth asphyxia, primarily due to its potential to cause fatal arrhythmias^12, 15^.

Hypocalcemia was strongly associated with convulsions, resuscitation, and prolonged intravenous fluid therapy. These associations mirror observations from Kenya and India, where seizures were a common presenting feature of hypocalcemia in neonates with asphyxia^5, 6, 12^. The mechanism likely involves impaired parathyroid hormone secretion and disruption of calcium homeostasis following hypoxic injury^5, 8^. In our study, hypocalcemia was the only electrolyte abnormality significantly associated with prolonged hospital stay, a finding consistent with reports from Bangladesh^12, 17^. This outcome may be explained by the recurrent seizures and neuromuscular instability that require extended monitoring and treatment in neonates with low calcium levels.

With regard to early outcomes, our study demonstrated that overall mortality was 11.8%, with the highest mortality occurring among neonates with hyperkalemia (41.4%). This finding is in agreement with Khondaker et al. in Bangladesh, who reported mortality of 60% among neonates with hyperkalemia compared to 22.6% among those with normal potassium levels^15^. The lethality of hyperkalemia lies in its direct effect on cardiac conduction, predisposing to arrhythmias and sudden cardiac death^9^. Hypocalcemia, on the other hand, did not significantly increase mortality but was associated with prolonged hospital stay in 48.4% of affected neonates compared to 18.6% among those with normal calcium. These results are comparable to findings from studies in India and Iraq, which showed that hypocalcemia contributes to morbidity by prolonging recovery rather than directly increasing mortality^12, 13^. Hyponatremia, though frequent, showed weaker associations with adverse outcomes in our study, a finding also echoed by some South Asian cohorts where sodium disturbances were more closely linked to illness severity than to death^11, 12^.

The clinical implications of our findings are important. Electrolyte derangements were frequent, often appeared early, and had distinct impacts on outcomes. Hyperkalemia was the strongest predictor of mortality, while hypocalcemia prolonged hospitalization. These results highlight the need for routine electrolyte monitoring as part of the standard management of neonates with birth asphyxia. Early detection and timely correction could reduce complications, shorten hospital stays, and prevent avoidable deaths. In settings such as Uganda, where routine electrolyte testing is not widely practiced due to resource limitations^3^, our findings provide strong evidence to advocate for inclusion of electrolyte monitoring in neonatal care protocols.

## Conclusion

This study demonstrated that electrolyte imbalances are common among term neonates admitted with birth asphyxia at Lira Regional Referral Hospital. Hyponatremia, hyperkalemia, and hypocalcemia were observed in nearly one-fifth to one-third of neonates, with most cases detected on day 3 of admission. Several clinical Predictors were independently associated with these disturbances, including low Apgar scores, severe hypoxic ischemic encephalopathy, convulsions, dehydration, resuscitation at birth, low birth weight, and prolonged intravenous fluid administration. Hyperkalemia was the strongest predictor of mortality, while hypocalcemia significantly prolonged hospital stay. These findings highlight the urgent need for routine monitoring of serum electrolytes in the management of asphyxiated neonates, as early detection and prompt correction can prevent complications and improve survival.

## Recommendations

All neonates admitted with birth asphyxia should be routinely screened for electrolyte imbalances, particularly sodium, potassium, and calcium, during the first days of hospitalization. In resource-limited settings where universal testing may not be feasible, priority should be given to neonates with severe hypoxic ischemic encephalopathy, low Apgar scores, those who required resuscitation, those with convulsions, low birth weight, dehydration, or those receiving intravenous fluids for more than 48 hours. Neonates found with electrolyte imbalances, especially hyperkalemia, should be closely monitored and promptly managed to reduce the risk of death. In addition, clinical protocols and guidelines on intravenous fluid use should be strengthened to minimize iatrogenic electrolyte disturbances. Strengthening routine monitoring of labour with the use of partographs is also essential to reduce the severity of asphyxia and its associated complications. Future research should explore long-term outcomes of electrolyte imbalance in birth asphyxia, including neurodevelopmental delay and cerebral palsy, which were beyond the scope of this study.

## Data Availability

The datasets used and/or analyzed during the current study are available from the corresponding author on reasonable request. Request should be sent to

## List of Abbreviations

APGAR: Appearance, Pulse, Grimace, Activity, Respiration
BA: Birth Asphyxia
HIE: Hypoxic Ischemic Encephalopathy
IV: Intravenous
NICU: Neonatal Intensive Care Unit
LRRH: Lira Regional Referral Hospital

## Ethics approval and consent to participate

Ethical approval was obtained from the Kampala International University Research Ethics Committee (Ref-NO-KIU-2024-755). Written informed consent was obtained from parents/guardians before enrollment.

## Consent for publication

Not applicable.

## Availability of data and materials

The datasets used and/or analyzed during the current study are available from the corresponding author on reasonable request. Request should be sent to (BY)

## Author contribution

**BY** was the principal investigator, conceived and designed the study, collected data, analysed data and wrote the draft of the manuscript. **TE, GN and SO** supervised the work and revised the manuscript, **ZIA, YAH, AAF, AMH, AO, HMY and TH** participated in data collection, revised the manuscript and all authors approved the final paper.

